# Results from EDIFICE : A French pilot study on COVID-19 and the gut microbiome in a hospital environment

**DOI:** 10.1101/2022.02.06.22269945

**Authors:** A.C.L. Cervino, R. Fabre, J. Plassais, G. Gbikpi-Benissan, E. Petat, E. Le Quellenec, L. Neuberger-Castillo, J-M. Laurent, L Iordache, M. Bouchahda, G. Marti, G. Chapelet

## Abstract

**BACKGROUND & AIMS:** Early reports suggest that both fecal shedding and dysbiosis of the gut microbiome are associated to disease severity in COVID-19 patients. We investigated the gut microbiome as well as the prevalence of SARS-CoV-2 in stool samples from two French populations: exposed healthcare workers and elderly hospitalized COVID-19 patients. The predictive power of bacterial loss of diversity and detection of SARS-CoV-2 in stool was assessed at 4 weeks against clinical outcomes in the patient group.

**METHODS:** 79 healthcare workers in contact with COVID-19 patients and 64 elderly patients hospitalised in a COVID-19 unit in France were included in the EDIFICE trial from April 2020 until May 2021. Stool samples were collected at inclusion. Loss of bacterial diversity was diagnosed based on 16S rRNA gene sequencing. Stool positivity to SARS-CoV-2 was determined by RT-PCR. Clinical outcomes were recorded at a 4 weeks follow up visit. In particular, these include whether the patient had been put under oxygen during the 4 weeks follow up, whether he had been discharged with or without aggravation from initial symptoms or whether the patient had died. The primary end point was to validate the hypothesis that hospitalized COVID-19 patients had more often lost their bacterial diversity than highly exposed active healthcare workers.

**RESULTS:** Elderly hospitalised patients with COVID-19 had more frequently lost their bacterial diversity when compared to exposed healthcare workers (p-value = 0.005), their severe dysbiosis was characterized by enrichment of the family Erysipelotrichaceae and depletion of beneficial bacteria at the genus level such as butyrate producers (Butyrivibrio, Roseburia, Faecalibacterium) and Bifidobacterium. The virus was detected in 61% of hospitalized patients and in only one healthcare workers (2%) who had previously been diagnosed with COVID-19 (p-value<0.001). No significant difference in the gut microbiome composition at the genus level of patients that tested positive in stool versus patients that tested negative was observed. Neither bacterial loss of diversity nor positivity to SARS-CoV-2 were associated to clinical outcome at 4 weeks.

**CONCLUSIONS:** We report findings of the first French trial investigating the clinical interest of stool based diagnosis of SARS-CoV-2 and loss of bacterial diversity in a population of elderly hospitalised COVID-19 patients and highly exposed healthcare workers. Our findings of reduced bacterial diversity and a strong gut dysbiosis in elderly hospitalized COVID-19 patients are highly consistent with previous reports mostly from Chinese populations. A major limitation is that observed differences in the gut microbiome between the two studied groups cannot be attributed to COVID-19 per se given the large number of confounding factors. SARS-CoV-2 was detected in the stool of the majority of hospitalized patients even several weeks after initial diagnosis by nasopharyngeal swabs. This high prevalence warrants further investigation by the scientific community into mechanism.

## INTRODUCTION

The ongoing Coronavirus Disease 2019 (COVID-19) pandemic, caused by the SARS-CoV-2 virus, is a worldwide pandemic. As of December 1^st^, 2021, more than 260□million confirmed cases and 5 million deaths have been reported (World Health Organization). In France, more than 7 million cases have been confirmed so far, resulting in over 110,000 deaths. Fortunately, the SARS-CoV-2 and its casualties have been partially controlled by implementation of nationwide sanitary measures, the widespread testing strategy and the fast development of vaccines. Interindividual contamination is thought to take place through inhalation of infected droplets or physical contact leading to the virus entering through the nose or the mouth. Patient diagnosis is mostly based on nasopharyngeal swabs. Fast and accurate diagnosis of the SARS-CoV-2 virus continues to take a central place in the pandemic control strategy and public health decision-making, with RT-PCR or antigenic tests but also with whole genome sequencing approach to understand transmission dynamics and follow new variants (Oude Munnink et al. 2020).

Main symptoms in hospitalized patients with mild to severe disease include cough, fever, fatigue and shortness of breath. Other symptoms are musculoskeletal, anosmia/dysgeusia and gastro-intestinal (GI) symptoms (Docherty et al. 2020)(D. Wang et al. 2020). Diarrhea is the most common GI symptom, reported in 2% to 50% of cases (Megyeri et al. 2021) mostly in association with respiratory symptoms (Docherty et al. 2020). Other GI symptoms include nausea, vomiting or abdominal pain in 2% to 10% of cases (Wong, Lui, and Sung 2020; D. Wang et al. 2020).

SARS-CoV-2 has been detected by RT-PCR in stool samples of COVID-19 patients (D. Wang et al. 2020; Xu et al. 2020; Wu et al. 2020; Cheung et al. 2020; Vaselli et al. 2021; Y. Zhou et al. 2021; Zuo et al. 2020; Britton et al. 2021). RT-PCR tests on faecal samples can remain positive about 50% longer than tests from nasopharyngal swabs after first symptom onset, meaning that the virus can be detected in stool samples even when it is no longer detectable by nasopharyngal swabs (Wu et al. 2020). On average stool-based detection can be positive for as long as 28 days raising the question of whether stool can be a source of continued contamination. (Wölfel et al. 2020) did however show that the virus is not infectious through that route despite elevated viral loads in a study on 9 patients. Other reports though indicate isolating the infectious virus from a stool sample (Yong Zhang et al. 2020; Xiao et al. 2020). Currently the infectious potential of virus titre found in faeces is still unclear (Guo et al. 2021).

Most research since 2019 has focused on the respiratory tract and to a much lesser extent has the gut been investigated. (Zuo et al. 2020) published the first cohort study investigating alterations in the gut microbiome and viral activity in stool. Since then, several other publications reported results on the differences between the gut bacterial microbiome in COVID-19 patients compared to controls (Gu et al. 2020; Chen et al. 2022; Mazzarelli et al. 2021; Yeoh et al. 2021; Reinold et al. 2021; Newsome et al. 2021; Tao et al. 2020; Li et al. 2021). Most reported a decreased bacterial diversity in the gut of COVID-19 patients when compared to controls. The observed dysbiosis was linked to enrichment of opportunistic pathogens and decrease of beneficial bacteria such as Butyrate producers and Bifidobacteria. Findings also support a possible association with disease severity (Tang et al. 2020; Zuo et al. 2020; Moreira-Rosário et al. 2021). The majority of studies were prospective cohorts from patients of Asian ethnicity, and the microbiome analysis was performed in a research environment. Here we report for the first time, the results from a prospective, observational, multicentric, controlled clinical trial (EDIFICE) using a commercial IVD test (1test1™) in a French population. It is also the first report on two distinct populations: highly exposed healthcare workers and elderly hospitalised patients.

## MATERIAL & METHODS

### Subject recruitment and sample collection

EDIFICE was approved by the French ethics committee, Comité de Protection des Personnes Est-III, under number 2020-A00979-30 on April 21^st^, 2020. All participants consented to the study. Study participants were recruited between April 28^th^, 2020, and April 29^th^, 2021, in one of the following four clinical centers: Clinique Saint-Jean L’hermitage (Melun, France), Clinique du Mousseau (Evry, France), Hôpital de Fontainebleau (Fontainebleau, France) and Centre Hospitalier Universitaire (CHU) de Nantes (Nantes, France). The study included two groups : a first group of healthcare workers working in a COVID-19 unit and representing an active population highly exposed to the virus through direct contact with COVID19 patients, and a patient group of elderly hospitalised patients treated in a COVID-19 unit. Study participants had to be between 18 and 95 years old and able to provide a stool sample non interventionally. Patients in critical care were therefore excluded. Study participants were all given at inclusion a commercial IVD home collection kit (1test1™, Luxia Scientific, France). Stool samples were thus collected using the DNA/RNA Shield™ Fecal Collection Tubes (Zymo Research, Irvine, California). Each sample contained approximately 1 gram of sample in 9 ml of stabilizer, representing a 1:10 dilution.

### Stool DNA Extraction

Samples for microbiome analysis were processed at the Integrated Biobank of Luxembourg (IBBL). All samples were processed within one month of stool collection following a validated stool DNA extraction protocol on the Chemagen MSM I instrument (PerkinElmer Chemagen, Baesweiler, Germany) (Mathay et al. 2015; Neuberger-Castillo et al. 2020) using the Chemagic™ DNA Blood 4k Kit special H24 with special lysis buffer for fecal samples. In brief, each sample of 1 ml was lysed by adding 2ml of SEB lysis buffer and 30µl Proteinase K, homogenized and incubated for 10min at 70°C, followed by 5min at 95°C. Lysates (1.5mL) were centrifuged for 5min at 10,000 g at RT. 1ml supernatants were transferred into a 24XL deep-well plate, which were processed using the magnetic beads based Chemagic™ MSM I automated protocol. Each run included bacterial DNA extraction controls (Bacterial Pool QC) in duplicates to check for the DNA extraction efficiency and possible contamination.

### DNA quantification and qualification

Total DNA quantification and purity (A260/A280 ratio) by spectrophotometry was performed using a Synergy Mx Monochromator-Based Multi-Mode microplate reader (Biotek) with Gen 5 version 2.0 software. Double-stranded DNA quantification by spectrofluorometry was performed with Quant-iT PicoGreen dsDNA Assay Kit (Invitrogen) using 96-well plates on the same device.

### 16S rRNA gene sequencing

The microbial composition profiling was performed by sequencing the V3-V4 regions of the prokaryotic 16S rRNA gene on the Illumina MiSeq using 2×300 bp paired-end reads. Amplicons were generated, cleaned, indexed and sequenced according to the Illumina-demonstrated 16S Metagenomic Sequencing Library Preparation Protocol with certain modifications (Illumina). In brief, an initial PCR reaction contained at least 12.5 ng of DNA.

A subsequent limited-cycle amplification step was performed to add multiplexing indices and Illumina sequencing adapters. Libraries were normalized and pooled, and sequenced on the Illumina MiSeq instrument.

Each library preparation run included an internal bacterial DNA extraction control (Bacterial Pool QC) and an internal 16S mock bacterial community control (DNA QC 16S) to detect any contamination during the sequencing library preparation and/or artefacts arising from the bioinformatics analysis. DNase/RNase-free water was used as negative control to detect any contamination. QC checks on fragment size, cluster density, error rate and Q30 were performed systematically all throughout the processing. Raw sequence data generated for this study are available in the Sequence Read Archive under BioProject accession PRJNA787810.

### Stool RNA extraction and RT-PCR

RNA was extracted from 250µl of sample using the ZymoBIOMICS™ RNA Miniprep kit (ZR2001) (Zymo Research, Irvine CA, USA). Validation work on Acrometrix™ COVID-19 control RNA (Thermo Fisher Scientific, Fremont CA, USA) indicated that high levels of inhibition persisted. To address this issue, an optimal dilution was established at 1:4 and an additional cleaning step with the Qiagen RNeasy MinElute Cleanup Kit was necessary (Qiagen, Hilden, Germany). RT-PCR was performed on a LightCycler (Roche, Vienna, Autria) using the Quick SARS-CoV-2 Multiplex Kit (CE-IVD, ref ZR3013, Zymo Research). All runs included a positive and negative control. A CP value <40 was considered positive.

### Bioinformatics and statistical analysis

Fastq files were analysed with 1TEST1SOFT, an internal bioinformatics pipeline that is based on Qiime2 (Bolyen et al. 2019) (2018.6) and RDP (Cole et al. 2014) (trainset n°16) and that has been extensively validated as part of the IVD status of 1test1™. The pipeline first removes expected 5’ and 3’ primers, CCTACGGGNGGCWGCAG and GACTACHVGGGTATCTAATCC, respectively, from the paired-end reads in the Fastq files, using the Cutadapt tool. Joined sequences are then quality-filtered using the Quality-filter plugin of Qiime2 with default parameters. A denoising step follows using the Denoise-16S method of the Deblur plugin with a trimming option set to 400bp, which produces amplicon sequence variants (ASVs) from which diversity is estimated. Taxonomic quantification of the denoised samples is finally obtained through the Classify-sklearn method of the Feature-classifier plugin, using the default confidence value (0.7) and a classifier model trained on the V3-V4 region (extracted using the above primers) of the reference sequences from the RDP database. Alpha-diversity has been considered through the Shannon index calculated in the Core-metrics-phylogenetic pipeline of the Diversity plugin of Qiime2 with a cut-off value of 5.57 for diagnosing the low bacterial diversity at a rarefaction of 9 000 reads.

Statistical analysis was performed internally using (“R: The R Project for Statistical Computing” n.d.) (v 4.1.0) and (“RStudio | Open Source & Professional Software for Data Science Teams” n.d.) (v 1.4.1106) according to the Statistical Analysis Plan established prior to database lock. Patient data have been 100% monitored by an independent contract research organization (CRO). For microbiome analysis a Wilcoxon-Mann-Whitney (WMW) test was performed on percent values for bacterial genera present in at least 70% in either group. P-values were adjusted for multiple testing using Benjamini-Hochberg.

### Loss of bacterial diversity diagnosis

The cutoff of 5.57 for the Shannon index was previously established based on the first quartile of alpha-diversity in a healthy group of 29 west European residents. The cutoff was further validated in a real life cohort of 100 participants who self reported their health status as one of five categories (“Excellent “, “Very good “, “Good “, “Not so good “, “Poor “). The frequency in the Excellent and Very good groups was 25% versus 87% in the self reported poor health group (Figure 1). The technical repeatability of the diversity measure was estimated to be 10%.

**Figure 1:**
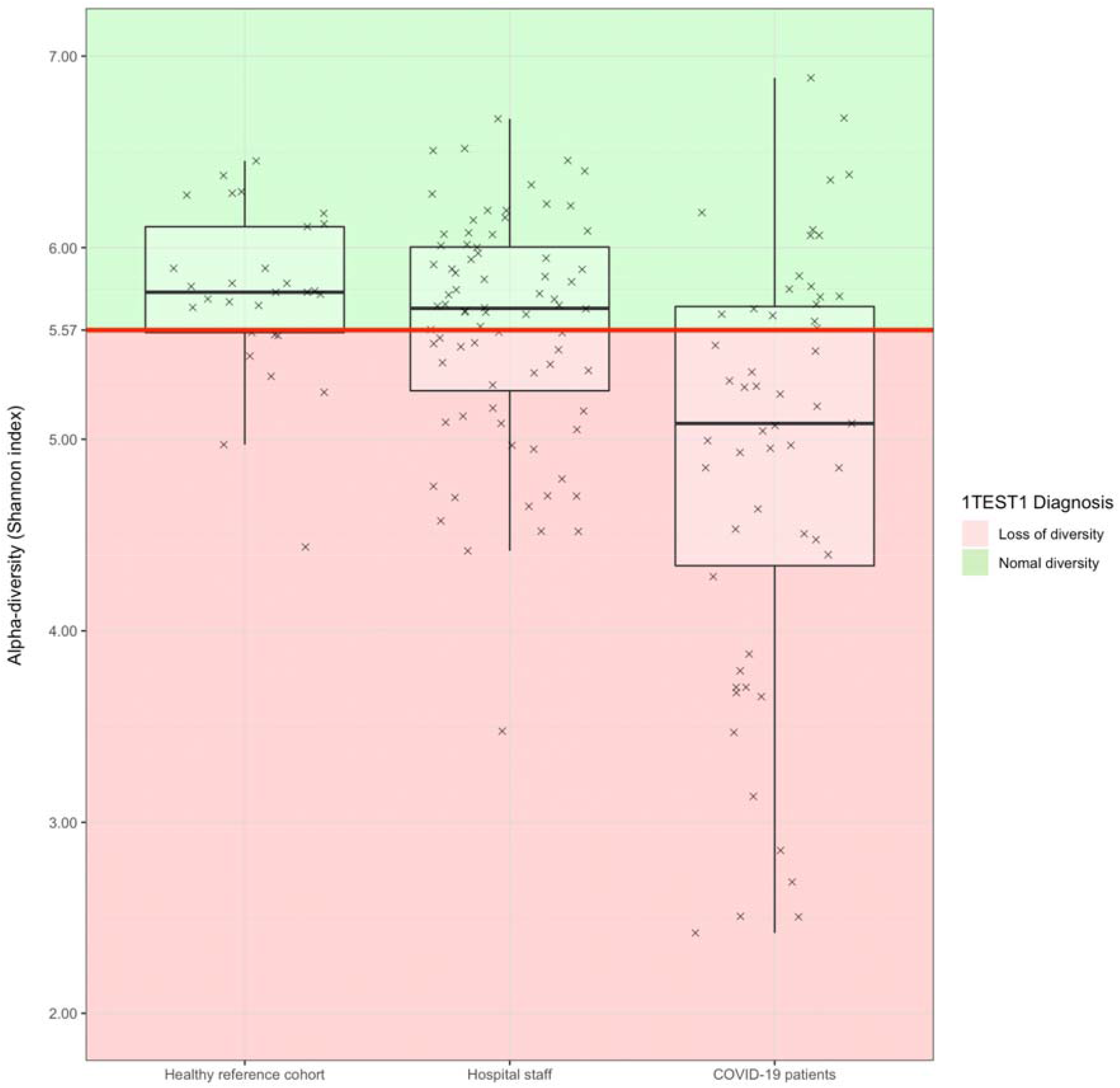
Alpha diversity values (crosses) and diagnosis of loss of bacterial diversity (red area) in the internal healthy reference cohort, and the two EDIFICE sub populations (Healthcare workers and COVID-19 Patients).

## RESULTS

### Clinical characteristics of the Study Population

In this trial we enrolled 143 participants: 64 elderly hospitalized COVID-19 patients and 79 healthcare workers working in close contact with COVID-19 patients. Amongst the patient group 62 had the follow up visit at 4 weeks. 55 patients had their microbiome successfully analysed by 16S rRNA gene sequencing and 61 were successfully analysed by RT-PCR for SARS-CoV-2 detection. 52 patients had results from both analyses (microbiome and viral detection). 24 of the 79 exposed staff self-reported having had COVID-19 like symptoms in the past but as those had not been confirmed by viral PCR tests, this variable was not analysed. None reported having had severe symptoms needing hospital care.

The two groups were unmatched and differed for most variables (Table 1). The majority of healthcare workers were nurses which in France are mostly women. This is reflected by 82% of Females in the healthcare workers population versus 61% in the COVID-19 hospitalised patient population. On average healthcare workers was 44 years younger than the COVID-19 patient population (41 y.o. versus 85 y.o.) Healthcare workers was also leaner (BMI of 23 versus 26). Variables such as age and BMI are known to be associated with alterations of the gut microbiome and in particular a lower alpha diversity. COVID-19 patients reported a higher rate hypercholesteremia (11%), Type II Diabetes (24%), Kidney failure (21%), history of stroke (13%) and immunodepression (16%). Most COVID-19 Patients had taken antibiotics (76%) in the previous 3 months as well as Proton Pump Inhibitors (89%) compared to a minority, respectively 9% and 5%, amongst the healthcare workers.

**Table 1:**
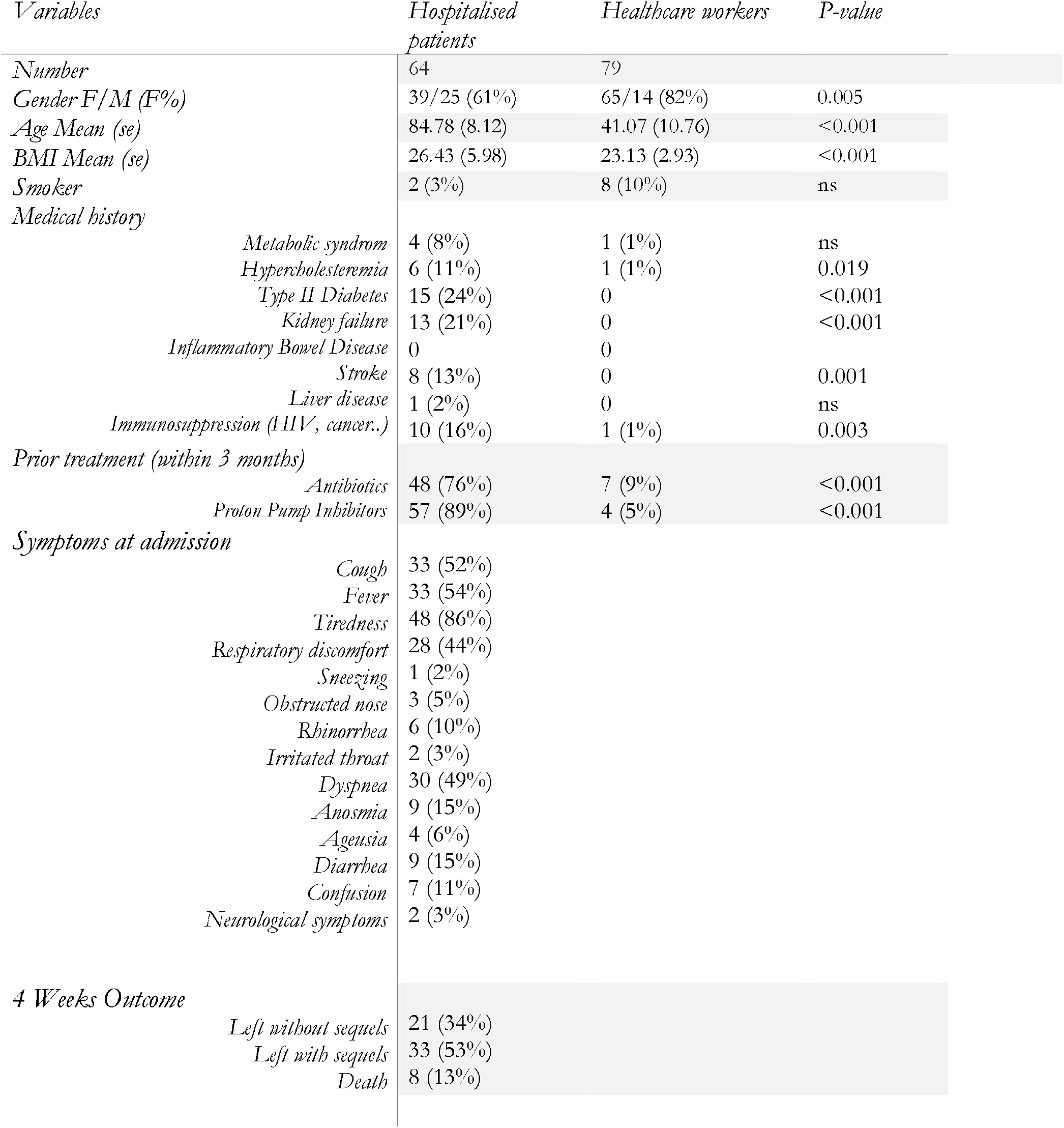
Demographic and clinical characteristics of the two groups (Elderly hospitalized COVID-19 patients and exposed healthcare workers).

In the COVID-19 population, the three most commonly reported symptoms at diagnosis were tiredness (86%), fever (54%) and cough (52%). Diarrhea was reported by 15% of patients.

### Loss of bacterial diversity and SARS-CoV-2 positivity

Loss of bacterial diversity was assessed in 76 healthcare workers and 55 elderly COVID-19 patients. 41% of the healthcare workers was diagnosed with a loss of bacterial diversity (Table 2a), which is slightly higher than the standard of 25% from the healthy reference European cohort used for calibration of 1test1™ (Figure 1). Amongst the elderly COVID-19 patients (Table 2a) 65% had lost their bacterial diversity which is an expected significant increase compared to the healthcare workers (p= 0.005, Chi-squared test).

**Table 2a:** Loss of bacterial diversity in the two groups (Elderly hospitalized COVID-19 patients and exposed healthcare workers)

| *Variables* | *Hospitalised*<br>*COVID-19*<br>*patients* | *Healthcare workers* | *P-value* |
| --- | --- | --- | --- |
| *Numbers* | 55 | 76 |  |
| *Loss of bacterial diversity* | 36 (65%) | 31 (41%) | 0.005 |

Amongst the healthcare workers only one person tested positive in stool, representing 2% of that population (Table 2b). Amongst the elderly COVID-19 hospitalised patients the majority tested positive (61%) which is an expected significant increase compared to the healthcare workers (p<0.001, Chi-squared test). Samples tested positive even 30 days after initial diagnosis (Supplementary Figure 1).

**Tableau 2b:** Stool based detection of SARS-CoV2 in the two groups (Elderly hospitalized COVID-19 patients and exposed healthcare workers).

| *Variables* | *Hospitalised*<br>*COVID-19*<br>*patients* | *Healthcare workers* | *P-value* |
| --- | --- | --- | --- |
| *Numbers* | 64 | 79 |  |
| *SARS-Co V-2 positivity* | 37 (61%) | 1 (2%) | <0.001 |

We also tested whether loss of bacterial diversity or presence of SARS-CoV-2 in stool was associated with the patient status 4 weeks after the stool collection. At 4 weeks, 34% of hospitalized SARS-CoV-2 patients had left the hospital without sequels, 53% had left the hospital with sequels, and 13% died (Table 1). Sequels at 4 weeks included loss of autonomy, undernutrition, oxygen supply and asthenia. None of those conditions were significantly associated with either loss of bacterial diversity (p-value=1 for death and p-value= 0.375 for without sequels) nor viral positivity in stool (p-value=0.138 for death and p-value= 0.404 for without sequels). 7 out of the 8 patients who died were positive for SARS-CoV-2 in stool (87.5%) however this was not statistically significant but should be confirmed in a larger cohort.

### Taxonomical comparison of the gut microbiome across different groups

Major differences were observed in the taxonomical composition between the hospitalised patients group and the healthcare workers. A total of 33 out of 56 predominant genera were found to be significantly altered in the gut microbiome (Table 3). 26 genera were decreased and 7 were increased in the hospitalised COVID-19 patients group versus the healthcare workers. Amongst the genera that are the most significantly decreased, one can find multiple beneficial bacteria such as the main butyrate producers (*Butyrivibrio. Roseburia and Faecalibacterium*) and *Bifidobacteria*. The seven bacteria that are increased in the elderly hospitalised COVID-19 patients are *Erysipelotrichaceae_Other, Eisenbergiella, Escherichia, Parabacteroides, Anaerotruncus, Streptococcus* and *Desulfovibrio*.

**Table 3:**
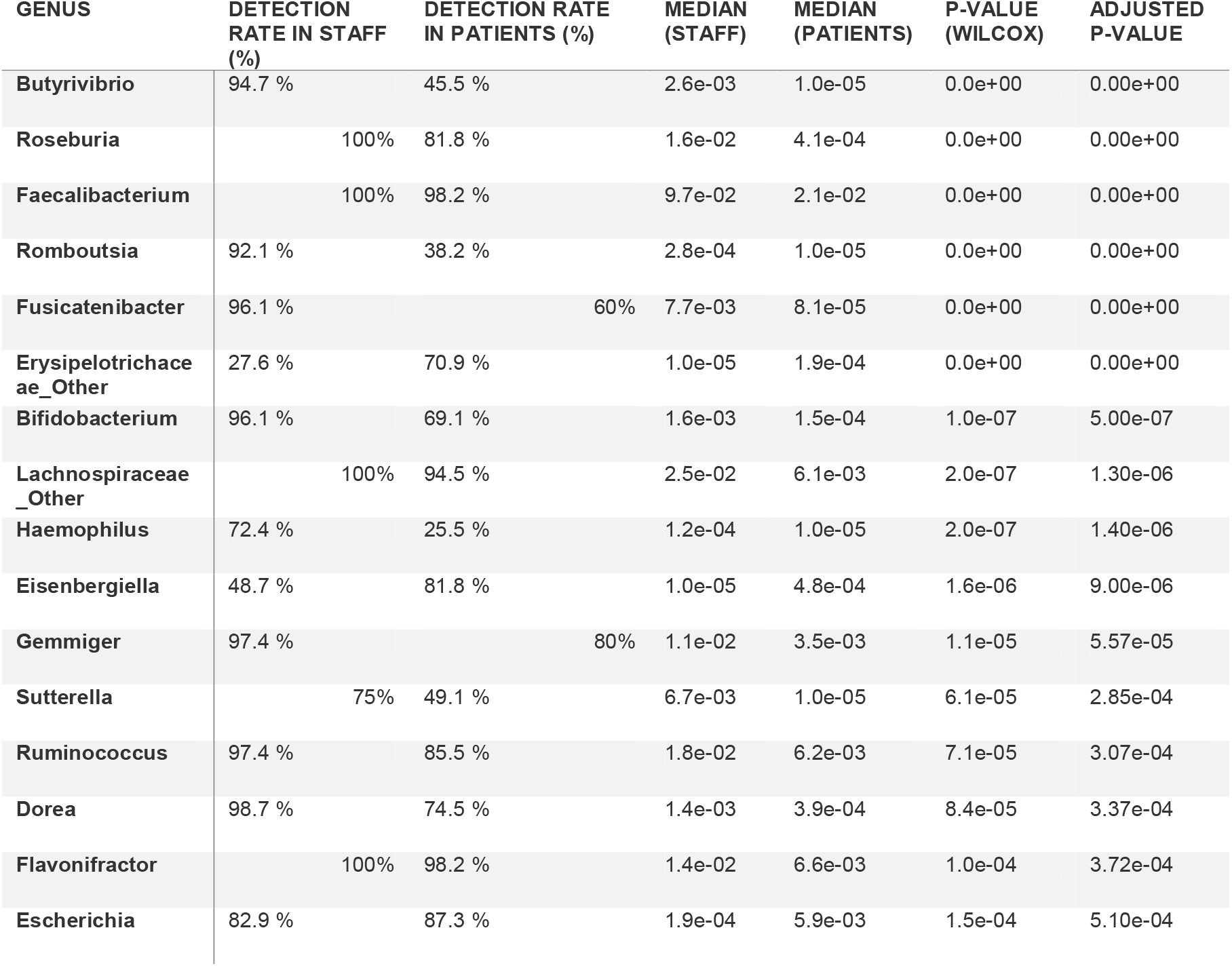
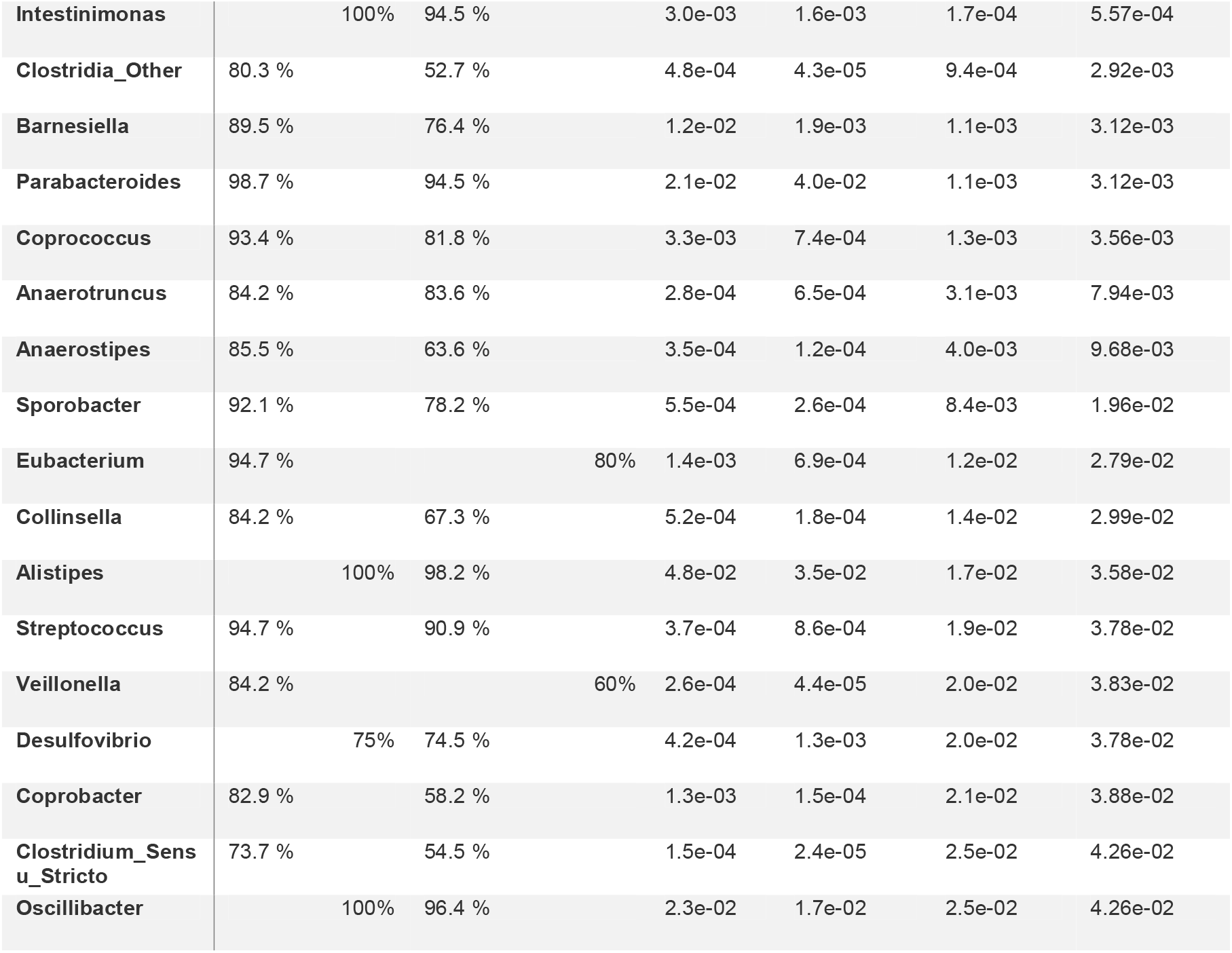
Taxonomical differences between the two groups (Elderly hospitalized COVID-19 patients compared to exposed healthcare workers).

**Table 4a:** Association between stool positivity and clinical outcome at 4 weeks.

| *Variables* | *Normal diversity*<br>*(N=19)* | *Loss of diversity*<br>*(N=36)* | *P-value* |
| --- | --- | --- | --- |
| *Death* | 2 (11%) | 5 (15%) | 1 |
| *Hospital leave without sequels* | 5 (26%) | 14 (41%) | 0.375 |

**Table 4b:**
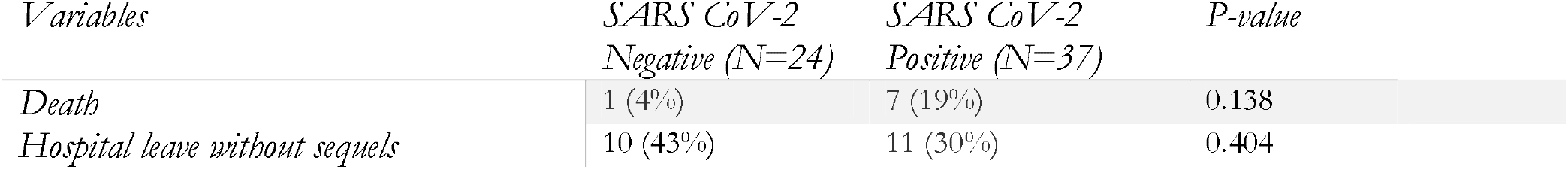
Association between stool positivity and clinical outcome at 4 weeks

On the other hand, no significant difference was observed in any of the genera when comparing the microbiome of patients that were SARS-CoV-2 positive (n=29) in stool to those with a SARS-CoV-2 negative result (n=23). Supplementary Table 1.

## DISCUSSION

We report for the first time on the clinical performance of a diagnostic test of loss of bacterial diversity, gut microbiome bacterial composition and stool positivity to SARS-CoV-2 in two distinct populations : a cohort of elderly hospitalized COVID-19 patients and a highly exposed active healthcare workers population.

Using the stool based self-collection kit 1test1™ (IVD, Luxia Scientific), participants were classified as either having a normal diversity or having a reduced diversity (also termed loss of diversity) based on a predefined cutoff value of the Shannon alpha diversity index. The initial motivation for a binary classification as “normal” and “lost diversity” in the general population is to identify people who would the most benefit from a dietary and life style change to improve their gut diversity. Using this binary classification of diversity, we found that a large proportion (65%) of elderly hospitalized COVID-19 patients had lost their bacterial diversity and that this value was significantly higher than our healthy reference cohort (25%) as well as the population of exposed healthcare workers (41%).

The reduced diversity in the patient group is consistent with previous reports indicating an overall lower alpha diversity in COVID-19 patients compared to controls ((Gu et al. 2020; Chen et al. 2022; Mazzarelli et al. 2021; Reinold et al. 2021; Tao et al. 2020; Moreira-Rosário et al. 2021; Li et al. 2021)), note that some authors reported on the Chao index instead of Shannon. Many environmental factors can contribute to a reduced alpha diversity such as urban life styles (Filippo et al. 2010), disease state (in particular metabolic syndrome (Le Chatelier et al. 2013) and IBD (Pascal et al. 2017)) and drugs (in particular antibiotics and PPIs) (Vich Vila et al. 2020) but not so much to age (Badal et al. 2020). Host genetics also impact gut microbial composition (Kurilshikov et al. 2021). The significant decrease in alpha diversity observed in elderly COVID-19 patients with significant co-morbidities was therefore expected and cannot, here, be attributed to COVID-19 status given the unmatched design of the trial. Previous reports indicate, however, that the decrease in alpha diversity is unlikely to be attributed to age or to antibiotics alone given that similar conclusions were reached in trials with matched designs or excluding previous antibiotics use (Gu et al. 2020; Reinold et al. 2021; Li et al. 2021) and that alpha diversity correlates to disease severity (Moreira-Rosário et al. 2021; Mazzarelli et al. 2021). In any case, evaluation within each group remains highly informative.

We identified a broad alteration of the gut microbiome composition between the two populations, with a significant difference in 33 out of the 56 predominant genera. The three most significantly decreased genera (*Butyrivibrio. Roseburia and Faecalibacterium)* are all butyrate producers. The significant decrease in butyrate producers (such as *Faecalibacterium, Roseburia, Eubacterium or Ruminococcus*) has consistently been reported by others on COVID-19 patients (Gu et al. 2020; Tang et al. 2020; Zuo et al. 2020; Mazzarelli et al. 2021; Yeoh et al. 2021; Reinold et al. 2021; Tao et al. 2020; Moreira-Rosário et al. 2021; Li et al. 2021) although not butyrivibrio specifically. Butyrate producers are well documented beneficial bacteria and are decreased in multiple diseases. Butyrate is an essential source of energy for colonocytes, improves the mucosal barrier and decreases inflammation (Leonel and Alvarez-Leite 2012). (F. Zhang et al. 2022) report decreased fecal butyrate levels in COVID19 patients and showed that this reduction is associated with increased plasma levels of inflammatory proteins. The observed decrease in SCFA and L-isoleucine biosynthesis in gut microbiome persisted even after recovery in patients. Bifidobacterium are also beneficial bacteria that are significantly decreased in COVID19 patients consistent with reports from others (Gu et al. 2020; Zuo et al. 2020; Yeoh et al. 2021; Reinold et al. 2021; Moreira-Rosário et al. 2021). Bifidobacterium have documented health benefits and many strains are used as probiotics.

Genera that are the most significantly increased in COVID19 patients, include Erysipelotrichaceae_Other, Eisenbergiella, Escherichia and Parabacteroides. Those genera have also been reported by others (Gu et al. 2020; Tang et al. 2020; Zuo et al. 2020; Mazzarelli et al. 2021; Yeoh et al. 2021; Reinold et al. 2021; Newsome et al. 2021; Li et al. 2021; Tao et al. 2020). The family Erysipelotrichaceae appears to be highly immunogenic, can potentially flourish post-treatment with broad spectrum antibiotics, and has been reported to be increased in colorectal cancer and metabolic disorders (Kaakoush 2015). This taxon does not in fact correspond to a genera, but rather a family level classification that could not be assigned at the genus level. Indeed, 16S based analysis does not allow accurate species level classification. Two other reports that used a shotgun metagenomic approach also reported Erysipelotrichaceae_noname as enriched in COVID-19 patients (Li et al. 2021) or associated with disease severity (Zuo et al. 2020). It could thus be interesting to perform de novo assembly of this bacteria in COVID19 patients to better characterize it. The genus Eisenbergiella has been reported to be correlated to cognitive impairment (Stadlbauer et al. 2020). Pro-inflammatory bacteria such as Escherichia are well known pathogens and are often reported in patients with diarrhea and in IBD cases.

The mechanism by which the virus enters the human host is becoming increasingly clear (Jackson et al. 2022). SARS-CoV-2 enters the host cells through binding top angiotensin-converting enzyme 2 (ACE2) receptors (P. Zhou et al. 2020; Q. Wang et al. 2020, 2) and the serine protease TMPRSS2 for S protein priming (Hoffmann et al. 2020). The ACE2 receptor is particularly highly expressed in colonic enterocytes (H. Zhang et al. 2020) (Y. Wang et al. 2020, 2). In mice, mutations in Ace2 leads to decreased expression of antimicrobial peptides and gut microbiome dysbiosis (Hashimoto et al. 2012). ACE2 is linked to amino acid transport in the gut (Cole-Jeffrey et al. 2015) and to tryptophan metabolism which is altered in COVID-19 patients (Gardinassi et al. 2020).

One of the main findings from our study is the large prevalence of SARS-CoV-2 observed in stool samples from hospitalized patients several weeks after initial infection (overall 61% reaching 87.5% in the subgroup of patients who died). For the exposed healthcare workers, the virus was only detected in one person that had been tested positive by nasopharyngal swab 35 days prior to sample collection and had not had any severe symptoms. Viral detection in fecal samples in 61% of COVID-19 cases is consistent with other reports that vary from 13% (Zhou 2021) to 73% (Zuo) with an estimated average of 40.3% from a meta-analysis conducted in 2020 (Parasa et al. 2020) and of 43% in 2021 (Yawen Zhang et al. 2021). SARS-CoV-2 is detectable in stool for a longer period than in nasophyringeal swabs, (Wu et al. 2020) estimate that on average samples stay positive for 27.9 days after the first symptoms later confirmed by a meta-analysis indicating 21.8 days (Yawen Zhang et al. 2021). Given the high rates of positivity in fecal samples, understanding the causes and consequences of SARS-CoV-2 presence in the digestive track are important issues that need to be addressed. Interestingly, persistence of SARS-CoV-2 nucleic acids detected in intestinal biopsies from subgroups of COVID19 patients 4 months post infection favorably impacts memory B cell response (Gaebler et al. 2021).

As pointed out by (Venzon and Cadwell 2022) “an important research direction would be to determine whether the gut microbiota dysbiosis observed in patients with severe COVID-19 affects the progression and recovery from the disease “. Here we investigated clinical outcome by looking at disease progression at 4 weeks, neither loss of diversity nor fecal viral RNA were associated with whether the patient had been put under oxygen during the 4 weeks follow up, whether he had been discharged with or without aggravation from initial symptoms or whether the patient had died. 7 out of the 8 COVID19 patients who died tested positive, although these results lacked statistical power, they are consistent with a previous report of association between death and fecal viral RNA (Britton et al. 2021). Analysis of clinical progression is important to assess the diagnostic value of biomarkers in a clinical setting. Although the four clinical outcomes we investigated here were not associated with loss of diversity or viral positivity, other clinical outcomes might be.

The EDIFICE trial suffers from various limitations, additionally to the lack of matching between groups, such as the lack of systematic testing of healthcare workers due to the non-interventional nature of the trial and it would have been interesting to systematically perform a follow up fecal RT-PCR test at 4 weeks in both groups. Additional exploration of inflammatory and antibody markers in blood and intestinal biopsies would also have been of great interest.

## Data Availability

Raw sequence data generated for this study are available in the Sequence Read Archive under BioProject accession PRJNA787810

https://www.ncbi.nlm.nih.gov/bioproject/787810

