## Supplemental Table 1 for "Results from EDIFICE : A French pilot study on COVID-19 and the gut microbiome in a hospital environment"

SUPPLEMENTARY TABLES & FIGURES

Supplementary Table 1 : Taxonomical differences between the two groups (Elderly hospitalized COVID-19 patients SARS-CoV-2 Negative in stool compared to elderly hospitalized COVID-19 patients SARS-CoV-2 Negative in stool).

| GENUS | DETECTION RATE IN PCR NEG PATIENTS (%) | DETECTION RATE IN PCR POS PATIENTS (%) | MEDIAN (PCR-POS) | MEdian (PCR-NEG) | p-value (Wilcox) | ADJUSTED p-value |
| --- | --- | --- | --- | --- | --- | --- |
| Phascolarctobacterium | 87 % | 69 % | 2.9e-02 | 1.4e-02 | 0.031 | 0.596 |
| Pseudoflavonifractor | 69.6 % | 93.1 % | 2.9e-04 | 5.5e-04 | 0.050 | 0.596 |
| Christensenella | 82.6 % | 93.1 % | 8.4e-04 | 2.0e-03 | 0.051 | 0.596 |
| Gemmiger | 82.6 % | 75.9 % | 6.4e-03 | 2.3e-03 | 0.054 | 0.596 |
| Ruminococcaceae_Other | 91.3 % | 96.6 % | 1.2e-02 | 3.0e-02 | 0.071 | 0.608 |
| Bilophila | 95.7 % | 79.3 % | 2.1e-03 | 1.4e-03 | 0.083 | 0.608 |
| Ruminococcus2 | 95.7 % | 93.1 % | 5.7e-03 | 1.6e-03 | 0.210 | 0.985 |
| Clostridium_IV | 95.7 % | 93.1 % | 2.2e-02 | 1.1e-02 | 0.347 | 0.985 |
| Parasutterella | 73.9 % | 65.5 % | 1.3e-03 | 2.4e-04 | 0.350 | 0.985 |
| Streptococcus | 91.3 % | 93.1 % | 7.2e-04 | 1.1e-03 | 0.357 | 0.985 |
| Faecalibacterium | 95.7 % | 100 % | 1.5e-02 | 2.1e-02 | 0.357 | 0.985 |
| Desulfovibrio | 69.6 % | 79.3 % | 1.3e-03 | 1.3e-03 | 0.403 | 0.985 |
| Flavonifractor | 95.7 % | 100 % | 8.4e-03 | 6.5e-03 | 0.484 | 0.985 |
| Roseburia | 78.3 % | 86.2 % | 2.4e-04 | 5.8e-04 | 0.494 | 0.985 |
| Eisenbergiella | 87 % | 79.3 % | 1.0e-03 | 2.5e-04 | 0.530 | 0.985 |
| Blautia | 95.7 % | 100 % | 3.8e-03 | 4.0e-03 | 0.543 | 0.985 |
| Bacteroides | 100 % | 96.6 % | 2.7e-01 | 2.4e-01 | 0.555 | 0.985 |
| Clostridiales_Other | 95.7 % | 96.6 % | 1.7e-02 | 3.1e-02 | 0.574 | 0.985 |
| Lachnospiraceae_Other | 95.7 % | 93.1 % | 5.3e-03 | 6.9e-03 | 0.580 | 0.985 |
| Dorea | 82.6 % | 69 % | 4.6e-04 | 3.5e-04 | 0.590 | 0.985 |
| Parabacteroides | 95.7 % | 93.1 % | 3.1e-02 | 4.2e-02 | 0.593 | 0.985 |
| Clostridium_XlVb | 78.3 % | 82.8 % | 2.1e-03 | 1.2e-03 | 0.598 | 0.985 |
| Barnesiella | 78.3 % | 72.4 % | 1.6e-03 | 2.7e-03 | 0.616 | 0.985 |
| Intestinimonas | 91.3 % | 96.6 % | 1.9e-03 | 1.4e-03 | 0.619 | 0.985 |
| Akkermansia | 73.9 % | 69 % | 7.6e-03 | 2.0e-03 | 0.654 | 0.985 |
| Escherichia | 91.3 % | 82.8 % | 5.9e-03 | 6.7e-03 | 0.658 | 0.985 |
| Lachnospiraceae_Other.1 | 95.7 % | 93.1 % | 2.5e-03 | 2.7e-03 | 0.672 | 0.985 |
| Clostridium_XVIII | 73.9 % | 75.9 % | 1.8e-04 | 1.7e-04 | 0.710 | 0.985 |
| Anaerotruncus | 87 % | 79.3 % | 6.8e-04 | 7.7e-04 | 0.726 | 0.985 |
| Eubacterium | 82.6 % | 75.9 % | 6.8e-04 | 7.2e-04 | 0.753 | 0.985 |
| Sporobacter | 73.9 % | 79.3 % | 2.6e-04 | 2.6e-04 | 0.753 | 0.985 |
| Coprococcus | 78.3 % | 82.8 % | 9.8e-04 | 4.1e-04 | 0.774 | 0.985 |
| Erysipelotrichaceae_Other | 82.6 % | 62.1 % | 1.9e-04 | 1.8e-04 | 0.794 | 0.985 |
| Hungatella | 73.9 % | 62.1 % | 1.8e-04 | 1.8e-04 | 0.822 | 0.985 |
| Holdemania | 73.9 % | 69 % | 1.3e-04 | 1.1e-04 | 0.837 | 0.985 |
| Alistipes | 100 % | 96.6 % | 3.5e-02 | 3.0e-02 | 0.854 | 0.985 |
| Odoribacter | 78.3 % | 75.9 % | 3.2e-03 | 2.9e-03 | 0.889 | 0.985 |
| Oscillibacter | 91.3 % | 100 % | 1.7e-02 | 1.7e-02 | 0.897 | 0.985 |
| Clostridium_XlVa | 100 % | 100 % | 3.5e-02 | 3.5e-02 | 0.912 | 0.985 |
| Ruminococcus | 87 % | 82.8 % | 3.8e-03 | 6.2e-03 | 0.948 | 0.985 |
| Prevotella | 78.3 % | 58.6 % | 1.1e-04 | 5.1e-04 | 0.955 | 0.985 |
| Butyricimonas | 73.9 % | 69 % | 1.1e-03 | 2.5e-03 | 0.970 | 0.985 |
| Butyricicoccus | 91.3 % | 93.1 % | 7.7e-04 | 1.3e-03 | 0.971 | 0.985 |
| Bifidobacterium | 73.9 % | 65.5 % | 1.2e-04 | 1.7e-04 | 0.985 | 0.985 |

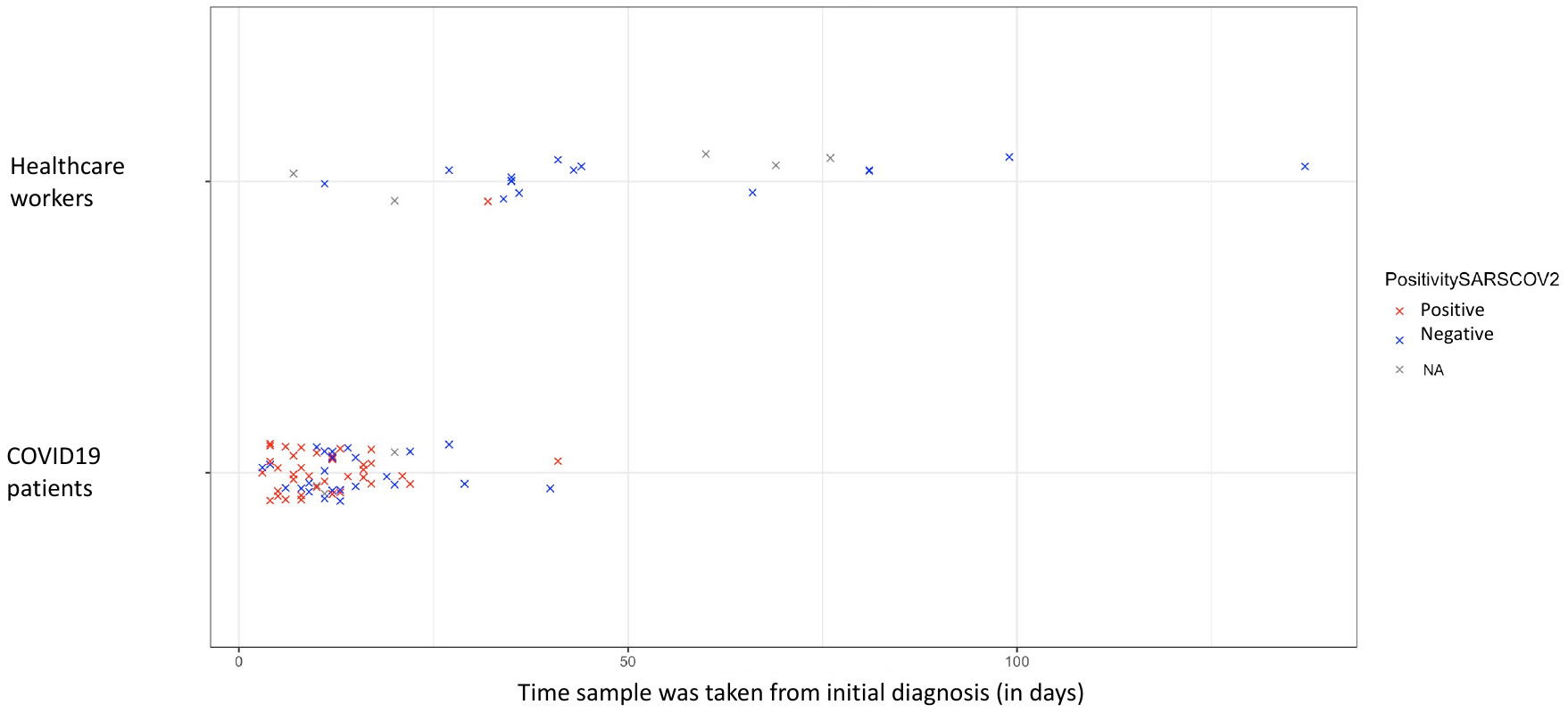

Supplementary Figure 1 : Stool based positivity to SARS-CoV-2 by collection time in the two groups
